# Multi-task deep autoencoder to predict Alzheimer’s disease progression using temporal DNA methylation data in peripheral blood

**DOI:** 10.1101/2022.04.02.22273346

**Authors:** Li Chen

## Abstract

**Motivation:** Traditional approaches for diagnosing Alzheimer’s disease (AD) such as brain imaging and cerebrospinal fluid are invasive and expensive. It is desirable to develop a useful diagnostic tool by exploiting biomarkers obtained from peripheral tissues due to their noninvasive and easily accessible characteristics. However, the capacity of using DNA methylation data in peripheral blood for predicting AD progression is rarely known. It is also challenging to develop an efficient prediction model considering the complex and high-dimensional DNA methylation data in a longitudinal study.

**Results:** We develop two multi-task deep autoencoders, which are based on convolutional autoencoder and long short-term memory autoencoder to learn the compressed feature representations by jointly minimizing the reconstruction error and maximizing the prediction accuracy. By benchmarking on longitudinal methylation data collected from peripheral blood in Alzheimer’s Disease Neuroimaging Initiative, we demonstrate that the multi-task deep autoencoders outperform state-of-the-art machine learning approaches for both predicting AD progression and reconstructing the temporal methylation profiles. In addition, the proposed multi-task deep autoencoders can predict AD progression accurately using only historical data and the performance is further improved by including all temporal data.

**Availability:** https://github.com/lichen-lab/MTAE

## 1 Introduction

The pathology of Alzheimer’s disease (AD) consists of amyloid-*β* (A*β*) deposition in the brain, the hyper-phosphorylation of tau proteins, and neuroinflammation through glial activation. Accordingly, brain imaging and liquid biopsy are the two main approaches for antemortem diagnosis of AD. Common diagnostic brain imaging tools include structural magnetic resonance imaging (MRI), functional MRI and positron emission tomography (PET). Especially, amyloid-PET is able to detect aggregated A*β* in the brain. Despite the success of imaging biomarkers in clinical practice, the economic burden and involved radioactive agents impede their widespread use in identifying AD. Compared to MRI and PET scans, fluid biomarkers in cerebrospinal fluid (CSF) are more accessible and affordable. However, spinal fluid tests require lumbar puncture, which are invasive and discomfort [13].

Due to the drawbacks of traditional ways for AD diagnosis, genetic biomarkers obtained from peripheral tissues such as blood would be a useful diagnostic tool due to its noninvasive and easily accessible characteristics. Moreover, emerging evidence suggests that epigenetics plays a significant role in AD pathogenesis, progression and resilience [26, 7, 18]. Particularly, DNA methylation is the most widely studied epigenetic mechanism for its ability to mediate gene expression. Accumulating evidence indicate that abnormal methylation can be used for detection and diagnosis of AD [31, 8]. Due to the attractive potentials of DNA methylation, multiple studies have been carried out to utilize DNA methylation data as predictors for AD diagnosis. For example, Bahado-Singh et al. adopts machine learning algorithms such as deep neural network and random forest to classify late-onset AD (LOAD) from cognitively healthy controls [2]. Park et al. utilizes a deep neural network to classify AD from controls by integrating both DNA methylation data and gene expression data [21]. However, none of these studies are longitudinal, so they can neither explore the dynamics of DNA methylation nor identify the converters switch between different disease stages.

The exponential reduction in sequencing cost, such as Illumina Infinium HumanMethylation450 Bead-Chip array (450K) or MethylationEPIC array (EPIC), increases the popularity of large-scale longitudinal methylation data, which facilitates the studies for methylation dynamics and disease progression such as bipolar disorder and type1 diabetes [3, 11]. Especially, Alzheimer’s Disease Neuroimaging Initiative (ADNI) recently provides a large cohort with longitudinal DNA methylation data collected from blood samples across different diagnostic groups such as cognitive normal (CN), mild cognitive impairment (MCI) and dementia (AD) [29, 28]. These DNA methylation data starts at baseline and leveraged up to 4 years of longitudinal DNA methylation data, sampled at approximately 1 year intervals to track the dynamic change of the methylation levels among the different diagnostic groups in the course of disease progression. This invaluable resource provide an unprecedented opportunity to understand how peripheral DNA methylation dynamics correlate with the diagnosis and progression of AD and to predict subjects who are more susceptible to AD progression.

However, existing work, for analyzing ADNI methylation data, mainly focus on the association analysis to identify a small set of significant epigenetic biomarkers associated with AD at baseline and progression [6, 15, 16]. The identified small number of epigenetic biomarkers can be potentially useful as a diagnostic tool due to the noninvasive nature and easily measurable characteristics. Besides the association analysis, another strategy for exploiting the longitudinal methylation data is to predict the converters, who are more easily converted from a mild to a more severe AD stage. However, developing computational methods to predict AD progression using longitudinal DNA methylation data comes with some challenges. First, different from epigenome-wide association studies (EWAS) to identify a small set of epigenetic markers with a strong effect size, predictive modeling may need to involve a much larger set of epigenetic markers with a moderate effect size to improve the prediction performance. Second, a large set of epigenetic markers results in a high-dimensional prediction task, which is a persistent challenge in the machining learning field. Third, longitudinal methylation data are nonlinearly correlated spatially and temporally. Yet, few studies has even been performed to predict AD progression using temporal DNA methylation data.

Deep learning approaches have recently been developed for AD diagnosis and progression by using the longitudinal data from various sources. Structural MRI has been used by convolutional neural networks (CNN) to classify the transition from the progression of MCI to AD and extract non-invasive MRI biomarkers linked to AD progression [17]. Recurrent neural network (RNN) and Long Short-Term Memory (LSTM) network have been adopted to predict AD progression by using a combination of features ranging from subject demographics, health history, clinical diagnosis, neuropsychological test scores, imaging markers and CSF measurements [30, 20, 12]. Recently, a combination of CNN and LSTM deep learning architecture (CNNLSTM) has been proposed to predict AD progression using structural MRI, where a CNN extracts the spatial features from images of multiple time points and a LSTM network learns the temporal dependency among extracted features from CNN for the prediction [5]. Different from clinical data and imaging data, the methylation data is high-dimensional. There are millions of CpGs on human genome compared to dozes of clinical features and brain images with a three-dimensional grid of dozens or hundreds voxels. To solve the challenge, recent work adopts a two-step approach for the high-dimensional methylation prediction by utilizing the advantage of both unsupervised and supervised deep learning architecture. First, an unsupervised deep learning model such as autoencoder (AE) has been introduced to learn compressed feature representations of high-dimensional methylation features. Second, the compressed feature representations are used as the model input for a supervised machine learning classifier such as SVM and deep neural network [9, 14]. However, none of these work adopt autoencoder to learn the compressed feature representations from longitudinal methylation data.

Despite the advantage of AE, it faces a limitation in a longitudinal study. AE needs to be performed on each time step to obtain the feature representations in each time step independently, which will ignore the temporal correlation of features. Recently, a novel autoencoder named convolutional autoencoder (CAE), which has been first introduced to denoise image data, has been used for time-series classification [23]. CAE can learn feature representations by jointly considering the spatial and temporal correlation of features. Then, the learnt feature representations can be used as model input for a classifier such as LSTM network [33]. Technically, the main difference between CAE and AE lies on the network structure for encoder and decoder. Different from fully-connected layers in both encoder and decoder in AE, CAE adopts convolution and transposed convolution operations for encoder and decoder respectively. Specifically, the temporal data can be structured in a 2D-matrix with time step and feature as two dimensions. In the encoder, a series of convolution operations can be performed to scan along the time dimension. In the decoder, a series of transposed convolution operations will reconstruct the temporal data. Another popular autoencoder for time-series classification is LSTM autoencoder (LSTMAE), The encoder consists of stacked LSTM layers to learn the high-order nonlinear temporal dependency of features. Each LSTM layer has the same number of LSTM units, and features in each time step connects to one LSTM unit in the first LSTM layer. The output of last LSTM unit in last LSTM layer is considered as the learnt feature representations, which is treated as the input to the decoder. The decoder reconstructs input features for each time step [10, 25, 32]. However, existing work for utilizing CAE and LSTMAE for time-series classification is still based on a two-step approach, where feature extraction using autoencoder is performed first and prediction using extracted features is carried out afterwards. Nevertheless, joint modeling feature extraction and prediction simultaneously may improve the performance for each task due to the benefit of multi-task learning.

Multi-task learning (MTL) is a field of machine learning, where multiple tasks are simultaneously learned by a shared model. MTL offers several advantages compared to single-task learning such as reducing overfitting through shared representations, and fast learning by leveraging auxiliary information [4]. However, despite these advantages of MTL, to the best of our knowledge, using MTL to predict AD progression using longitudinal DNA methylation data is never exploited. To fill this gap, we present two multi-task deep autoencoders with the primary task for predicting AD progression and secondary task for reconstructing the input methylation data simultaneously by utilizing longitudinal DNA methylation data collected from peripheral blood in Alzheimer’s Disease Neuroimaging Initiative (ADNI). Particularly, we propose <u>M</u>ulti-<u>T</u>ask <u>L</u>ong <u>S</u>hort-<u>T</u>erm <u>M</u>emory <u>A</u>uto<u>E</u>ncoder (MT-LSTMAE) and <u>M</u>ulti <u>T</u>ask <u>C</u>onvolutional <u>A</u>uto<u>E</u>ncoder (MT-CAE). MT-LSTMAE takes advantage of LSTM’s capacity for learning the temporal dependency on DNA methylation data in a longitudinal study. Similarly, MT-CAE leverages convolution operations to learn both spatial and temporal dependency of DNA methylation data. We further develop a hybrid loss function, which is a weighted average of the prediction error and reconstruction error. By minimizing the hybrid loss function, the multi-task deep autoencoders try to improve the learning for compressed feature representations of methylation data, and improve the prediction performance of the classifier using the compressed feature representation at the same time. We further benchmark two proposed multi-task models against existing deep learning methods for time-series classification such as CNN, LSTM and a hybrid of CNN and LSTM, as well as the two-step approach to use autoencoder for dimension reduction first and an independent classifier for prediction afterwards. Consequently, we find that MT-CAE and MT-LSTMAE significantly outperform competing methods in predicting CN-to-MCI and MCI-to-AD conversions especially when the beta values are unscaled. Moreover, we validate multi-task learning is superior to single-task learning by comparing MT-CAE and MT-LSTMAE to their counterparts with the same network architecture of autoencoder as well as the two-step approach. Furthermore, we demonstrate that using historical data only can accurately predict AD progression, and utilizing temporal methylation data can further improve the prediction. Lastly, we show that multi-task deep autoencoders achieve better performance in reconstructing the temporal methylation data than a standard autoencoder especially when beta values are unscaled. Altogether, we believe that the proposed deep learning approaches will benefit the AD diagnosis by using high-dimensional multi-omics profiles collected from peripheral blood in a longitudinal study.

## 2 Methods

### 2.1 Longitudinal DNA methylation data in ADNI

We download1905 DNA methylation samples collected from peripheral blood in a cohort of 649 unique individuals, who have participated in ADNIGO and ADNI2 in Alzheimer’s Disease Neuroimaging Initiative (ADNI). These DNA samples are longitudinal, which starts at baseline and leveraged up to four more visits with an approximate one year interval to model the alteration in methylation levels to delineate methylation dynamics associated with aging and disease progression. Each DNA methylation sample is profiled by Illumina Infinium HumanMethylationEPIC BeadChip Array, which covers ∼866,000 CpGs signals. Samples were randomized using a modified incomplete balanced block design to match age and sex and avoid confounding.

We use two R/Bioconductor packages IlluminaHumanMethylationEPICmanifest and IlluminaHuman-MethylationEPICanno.ilm10b4.hg19 to process the raw methylation data. Specifically, we use read.metharray.exp, preprocessRaw, ratioConvert, mapToGenome and getBeta functions to obtain the beta values with a value between 0 and 1 for each CpG. We then use ChAMP pipeline [19] for normalization, batch correction and cell type correction of CpG signals, and CpGs signals of biological replicates are averaged. As a result, we obtained the normalized methylation profiles for all CpGs in each individual.

Based on the clinical data, we observe three types of AD diagnosis: cognitive normal (CN), mild cognitive impairment (MCI) and dementia (AD). Among the 649 individuals, there are 221 CN, 334 MCI and 94 AD at the baseline visit. However, the distribution of AD diagnosis changes to 177 CN, 259 MCI and 213 AD at the last visit of each individual, which indicates an overall AD progression in the longitudinal study. The goal of the study is to evaluate the practicability and feasibility of using temporal DNA methylation data collected from peripheral blood for predicting different types of AD conversion.

To construct the positive set, we start from counting number of diagnosis for each individual in the longitudinal course and we are interested in individuals with the change of diagnosis. As a result, we identify 474 individuals have no change of diagnosis, 167 individuals has 2 different diagnosis and 8 individuals has 3 different diagnosis. Among 167 individuals with 2 different diagnosis, we find that majority of the individuals either changes from CN to MCI (n=44) or change from MCI to AD (n=111). Then, we use CN-to-MCI and MCI-to-AD respectively to construct two positive sets in the prediction task. We further construct the negative set using individuals with no change of diagnosis. Consequently, we identify 147 individuals with CN as baseline and no change (CN-to-CN) and 147 individuals with MCI as baseline and no change (MCI-to-MCI) during all visits. Since the number of visits are uneven and one diagnosis may be the same for several visits, we use the DNA methylation data at the first visit and the last visit for each individual as the temporal features in the positive set. Therefore, for each individual, there are two DNA methylation profiles in two time steps. Finally, we can develop computational models to classify CN-to-CN (n = 147) from CN-to-MCI (n = 44); and MCI-to-MCI (n = 147) from MCI-to-AD (n = 111) respectively.

### 2.2 Feature selection for the prediction task

To measure the methylation level, we use beta values, defined as 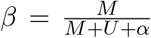, where *M* and *U* are the methylated and unmethylated signal intensities, and *α* is an offset. Since there are ∼866,000 CpGs, we need to perform a feature selection step to choose informative CpGs before fitting the prediction models. Here, we use a simple feature selection based on ratio of variance. Specifically, for *i*th CpG, we first calculate two variances of beta values in conversion group (e.g. CN-to-MCI) and non-conversion group (e.g. CN-to-CN) respectively, denoted as 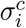 and 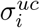. Second, we calculate the ratio of variance denoted as 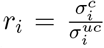. We then select the top 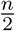 CpGs with largest ratios and bottom 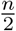 CpGs with smallest ratios. The *n* CpGs are considered as informative features. For a sensitivity analysis of *n* in predictive modeling, we exam the *n* from 1000, 2000 to 4000.

### 2.3 Architecture for multi-task deep autoencoder

Figure 1 illustrates an overview of our proposed two multi-task deep autoencoders to predict AD progression using DNA methylations data in a longitudinal study. The two autoencoders are Multi-Task Convolutional AutoEncoder (MT-CAE) (Fig 1A) and Multi-Task Long Short-Term Memory AutoEncoder (MT-LSTMAE) (Fig 1B). Either MT-CAE and MT-LSTMAE consists of an encoder and a decoder. The encoder aims to learn compressed feature representations that captures the spatial and temporal information from the temporal methylation profiles. The decoder reconstructs the temporal methylation profiles using compressed feature representations. In addition, the feature representations are used as the model input for a classifier to predict AD progression. In the following sections, we will illustrate the details of network architectures for MT-CAE and MT-LSTMAE respectively.

**Figure 1:**
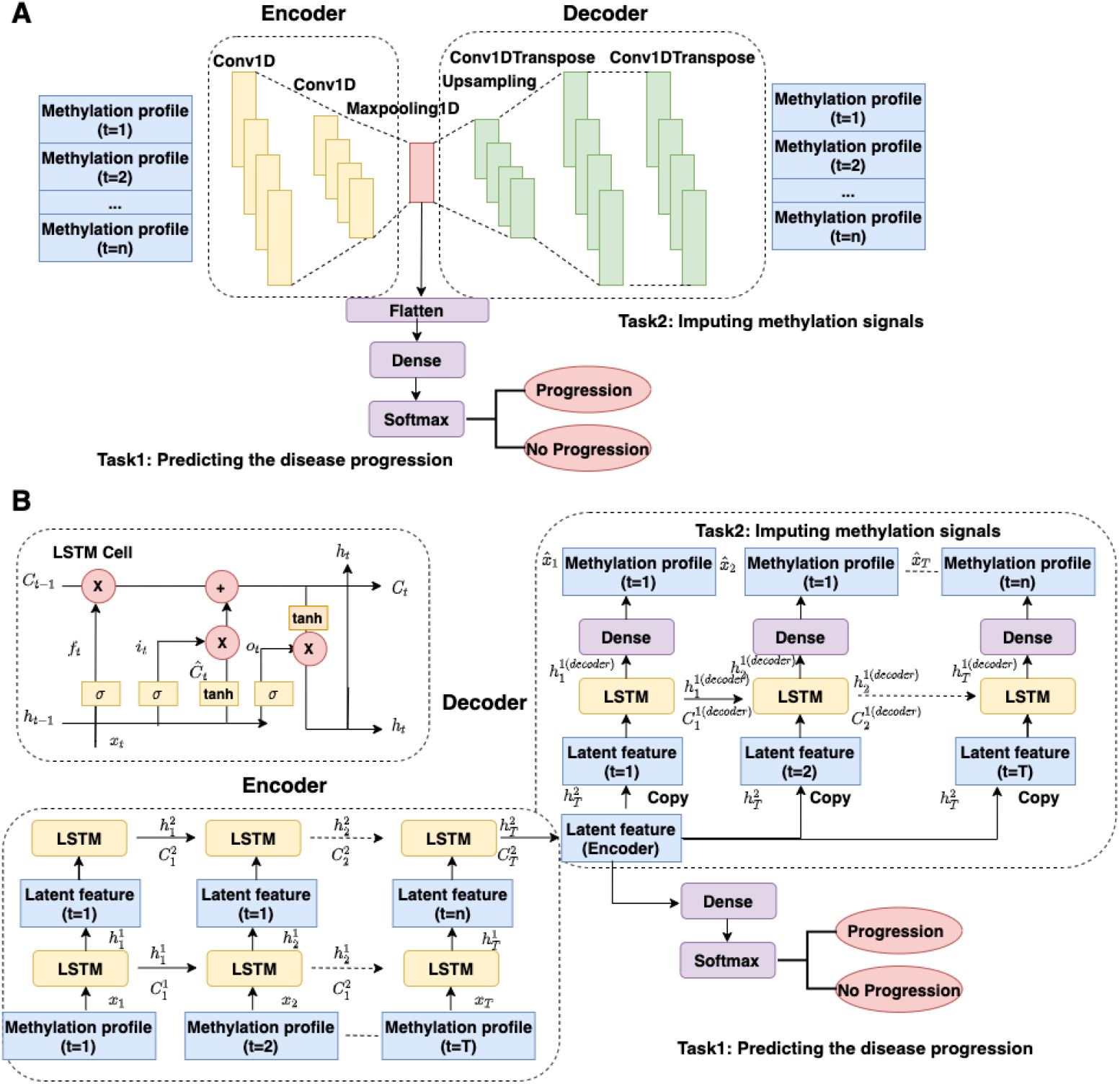
The flowchart for proposed two multi-task deep autoencoders. (A) Multi-task convolutional autoencoder (MT-CAE) (B) Multi-task LSTM autoencoder (MT-LSTMAE)

#### 2.3.1 Multi-task convolutional autoencoder

Similar to AE, CAE is an unsupervised dimensionality reduction model, which is composed by convolutional layers, to learn compressed feature representations. By minimizing reconstruction errors, CAE can remove noise from the images while simultaneously keeping useful information as much as possible to produce robust features. Different from AE, which requires the input image to be flattened into a single vector as feature input to the dense layers, CAE enjoys the benefit of working on the original image directly without disrupting the spatial information. The compressed feature representation can be used for the prediction task by connecting to a classifier [23].

Due to the advantage of CAE, we format the temporal DNA methylation data into a two-dimensional “image” format, which is formularized as 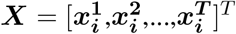, with time dimension *T* as row and feature dimension *p* as column. 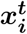 is a *p*-dimensional vector representing the methylation profile of a subject at time step *t*; *T* is the number of time steps. To perform the convolution, the convolution kernel is designed with the kernel size as *m* × *p* (*m* ≤ *T*). The convolution kernel moves along the time axis in one direction from the baseline to time *T* to perform convolution. In this way, convolution operation can capture both the spatial dependence on feature dimension and temporal dependency on time dimension.

In the encoder (Figure 1A), the methylation profiles undergo two convolutional layers to generate the feature map, which captures both spatial and temporal information from the temporal methylation data. Then, a maxpooling layer collapses the feature map into the one-dimensional feature representations ***z***. In the decoder, ***z*** is upsampling and goes through two transposed convolutional layers to reconstruct the methylation signals 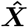. Overall, the encoder can be formularized as ***z*** = *Conv*_2_(*Conv*_1_(***X***)). The decoder can be formularized as 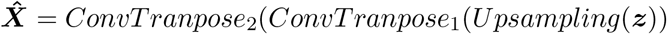. For the primary task, ***z*** is sent to to a feed-forward neural network that has a binary node with a softmax activation for predicting AD progression: 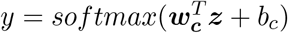, where ***w***_***c***_ and *b*_*c*_ are the weight and bias in the softmax layer. For the multi-task model, we develop a hybrid loss *L*_*T*_ as the total loss, which is a weighted average between the the prediction loss *L*_*P*_ and reconstruction loss *L*_*R*_. *L*_*P*_ is represented by the binary cross-entropy, which is formularized as,

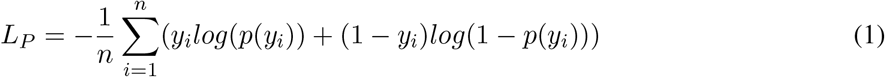

where *n* is the sample size for the batch; *y*_*i*_ is the label (1 for converter; 0 for non-converter) for *i*th sample and *p*(*y*_*i*_) is the predicted probability of *i*th sample being a converter.

*L*_*R*_ is measured by the mean squared error, which is formularized as,

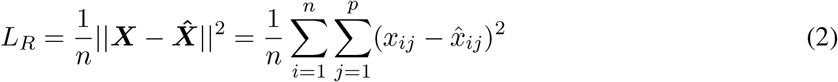

*L*_*T*_ is a weighted average between *L*_*P*_ and *L*_*R*_, which is formularized as,

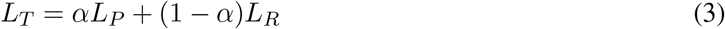

In practice, the weight *α* is treated as a hyper-parameter in the model selection.

#### 2.3.2 Multi-task LSTM autoencoder

First, we briefly introduce the concept and structure of LSTM network before integrating it into MT-LSTMAE. A LSTM network can contain multiple LSTM layers and each LSTM layer consists of multiple LSTM unit (Figure 1B). A common LSTM unit at time step *t* is composed of an input gate ***i***_*t*_, an output gate ***o***_*t*_ and a forget gate ***f***_*t*_. Regulated by the three gates, the LSTM unit is able to remove or add information to the cell state ***c***_*t*_. Specifically, the input for the forgotten gate ***f***_*t*_ includes the current input ***x***_*t*_, the hidden state ***h***_*t*_ and the previous hidden state ***h***_*t*−1_, which is activated by sigmoid function 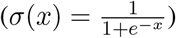,

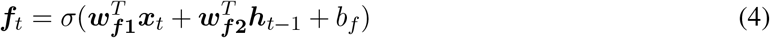

The input for the input gate ***i***_*t*_ is the same as that of the forget gate ***f***_*t*_ and further activated by sigmoid function,

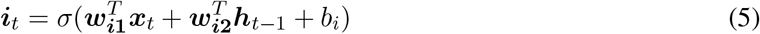

Cell state ***c***_*t*_ is an average of cell state at the previous time ***c***_*t*−1_ weighted by ***f***_*t*_ and a new candidate value 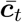 weighted by ***i***_*t*_. It should be noted that both ***f***_*t*_ and ***i***_*t*_ contains a value between 0 and 1 and are used as as weight for calculating cell state ***c***_*t*_.

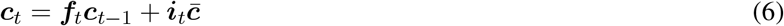

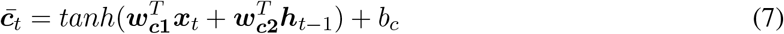

The input of the output gate ***o***_*t*_ is the same as that of input gate ***i***_*t*_ and the forget gate ***f***_*t*_,

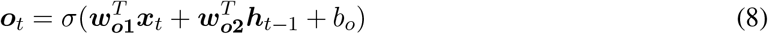

Finally, the hidden state ***h***_*t*_ is processed by tanh function 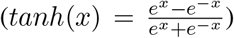 and multiplied by ***o***_*t*_, which is a value between 0 and 1 to control the information from ***c***_*t*_ to the hidden state ***h***_*t*_,

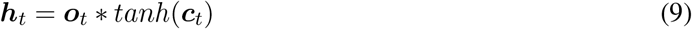

The new cell state ***c***_*t*_ and the hidden state ***h***_*t*_ at time step *t* inherits the historical information from the previous time step *t* − 1 and pass the current information to time step *t* + 1 to learn the temporal relationship. Moreover, in the above formulation, ***w***_***f*1**_ and ***w***_***f*2**_ are weights for forget gate; ***w***_***i*1**_ and ***w***_***i*2**_ are weights for input gate; ***w***_***o*1**_ and ***w***_***o*2**_ are weights for output gate; ***w***_***c*1**_ and ***w***_***c*2**_ are weights for cell state. *b*_*f*_, *b*_*i*_, *b*_*o*_ and *b*_*c*_ are the corresponding bias. All parameters are learnt in the model training.

Similar to MT-CAE, MT-LSTMAE also consists of an encoder to learn the feature representations and a decoder to reconstruct the temporal methylation data, as well as a classifier using the feature representations for prediction. Specifically, the encoder is composed of two-level stacked LSTM layers to learn the high-order nonlinear temporal dependency of methylation data. Each LSTM layer contains *T* LSTM units. The network structure and information flow are similar between the first and second LSTM Layer. In the first LSTM layer, *t*th LSTM unit takes the methylation profile at *t*th time step as the input. The first LSTM layer generates an output 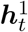 per time step as the input for *t*th LSTM unit in the second LSTM layer. The second LSTM layer has only one output 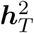 in the last time step *T* · 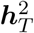 is also the output from the encoder, which is deemed as the compressed feature representations. Different from the encoder, the decoder consists of one LSTM layer with *T* LSTM units, and each LSTM unit is followed by a dense layer with shared weights across *T* time steps. The feature representation 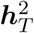 generated from the encoder is first repeated *T* times at the sequence input for *T* LSTM units in the LSTM layer in the decoder. The output of each LSTM unit at *t*th time step 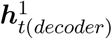 enters into a dense layer for predicting the methylation profiles at *t*th time step.

Similar to MT-CAE, feature representation 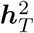 is flattened and entered into a feed-forward neural network that has a binary node with a softmax activation for predicting AD progression: 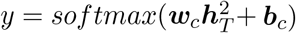, where ***w***_***c***_ and *b*_*c*_ are the weight and bias in the softmax layer. The prediction loss, reconstruction loss and final loss are defined the same as these of MT-CAE.

### 2.4 Competing machine learning methods

We compare the proposed two multi-task deep autoencoders to several classic deep learning approach (Table 1): (i) Convolutional neural networks with different number of convolutional layers (i.e. CNN1, CNN2), which have been widely used for time-series classification [34]. Similar to MT-CAE, the convolution operation is performed along the time axis; (ii) a simple one-layer LSTM network (i.e. LSTM1) and two-layer stacked LSTM network (i.e. LSTM2); (iii) a variation of LSTM named Bidirectional LSTM network with different number of Bidirectional layers (i.e. BiLSTM1, BiLSTM2), which can propagate the temporal information in both directions; (iii) a hybrid of CNN and LSTM network (i.e. CNNLSTM1, CNNLSTM2). The hybrid model can leverage CNN for feature extraction and LSTM network for learning the long-term temporal dependency; (iv) a common used practice for applying autoencoder in temporal data, which using a standard autoencoder to learn compressed features representations of high-dimensional input. The compressed features representations will be used as the model input for a classifier such as LSTM (i.e. AE-LSTM); (v) random forest, which is a robust non-deep learning approach for high-dimensional prediction [27]. However, only features at the last time step are used as the model input.

**Table 1:**
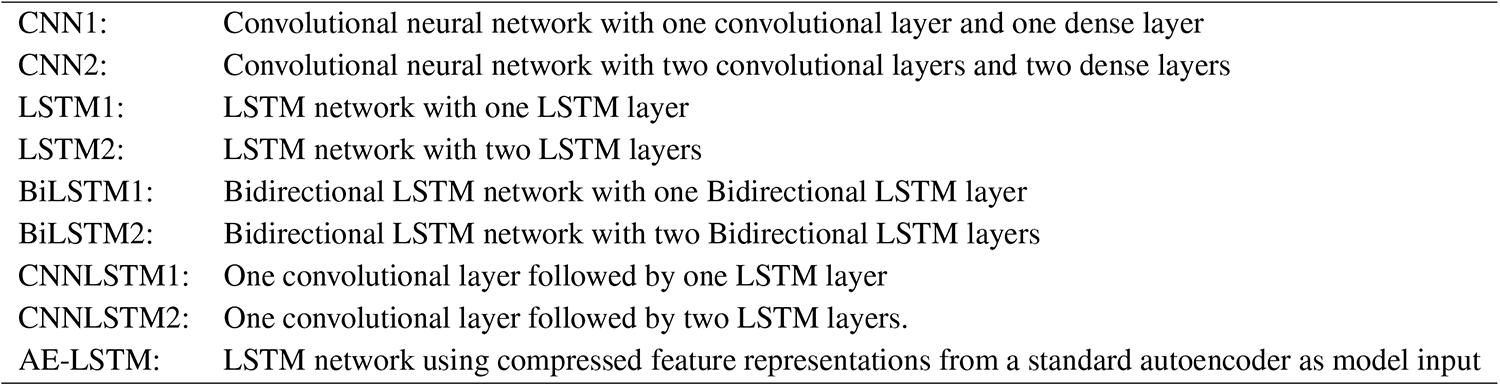
Competing deep learning methods

|  |  |
| --- | --- |
| CNN1: | Convolutional neural network with one convolutional layer and one dense layer |
| CNN2: | Convolutional neural network with two convolutional layers and two dense layers |
| LSTM1: | LSTM network with one LSTM layer |
| LSTM2: | LSTM network with two LSTM layers |
| BiLSTM1: | Bidirectional LSTM network with one Bidirectional LSTM layer |
| BiLSTM2: | Bidirectional LSTM network with two Bidirectional LSTM layers |
| CNNLSTM1: | One convolutional layer followed by one LSTM layer |
| CNNLSTM2: | One convolutional layer followed by two LSTM layers. |
| AE-LSTM: | LSTM network using compressed feature representations from a standard autoencoder as model input |

### 2.5 Model implementation, training, testing and hyperparameter tuning

We implement all deep learning models using Python Keras/Tensorflow v2 [1]. To further increase the training efficiency, we adopt “EarlyStopping” and “ModelCheckpoint” two techniques. EarlyStopping stops the training when prediction performance on validation set stops improving, which can happen before all epochs ends. Thus, EarlyStopping can help reduce training time. To stabilize EarlyStopping, we add a delay to trigger EarlyStopping by 5 epochs if there is no improvement of the prediction performance. The prediction performance is measured by validation loss for autoencoder and validation accuracy for other deep learning models. Together with EarlyStopping, ModelCheckpoint saves the trained model, whose prediction performance on validation set still improves. Therefore, the trained model with best prediction performance on the validation dataset is saved, which is not necessary the model trained in the last epoch.

For each prediction model, 20% of the total samples is used as independent testing, 80% is used as training set among which 20% is used as validation set. To control the bias from random sampling, the aforementioned procedure is repeated 50 times. We report the AUC and AUPRC of 50 experiments for each method and perform the method comparison using two-sided paired Wilcoxon rank-sum test.

For the hyperparameter tuning, the search space for number of LSTM units or number of convolutional kernels is a sequence with maximum number as half of the feature size and decreased by two-folds. The search space for dropouts is a sequence of (0.2,0.4,0.6,0.8). For multi-task deep autoencoders, the search space for the weight *α* of the prediction loss function is a sequence of (0.1,0.3,0.5,0.7,0.9). We adopt RandomResearch tuner from kerasTuner [24], which will automatically select the best combinations of hyperparameters in the search space. Specifically, evaluation metric is set as validation loss for autoencoder and validation accuracy for other deep learning models, and the number of maximum trials per combination of hyperparameters is set as 20. For random forest, we implement random forest with default parameters using the Python machine learning library “scikit-learn” [22].

## 3 Results

### 3.1 Prediction performance compared to existing machine learning methods

To demonstrate the advantage of multi-task deep autoencoders, we compare MT-CAE and MT-LSTMAE to eight common practices of using deep learning for temporal prediction, which mainly fall within three categories: (i) CNN with different number of convolutional layers (i.e. CNN1, CNN2); (ii) LSTM and Bidirectional LSTM with different number of LSTM layers (i.e. LSTM1, LSTM2, BiLSTM1, BiLSTM2) and (iii) a hybrid of CNN and LSTM with different number of LSTM layers (i.e. CNNLSTM1, CNNLSTM2). We benchmark all methods in predicting the two types of conversions: one is from cognitive normal (CN) to mild cognitive impairment (MCI), and the other is from mild cognitive impairment (MCI) to dementia (AD). For each conversion type and each prediction model, 20% of the total samples is used as independent testing, 80% is used as training set among which 20% is used as validation set. To control the bias from random sampling, the aforementioned procedure is repeated 50 times. We report the AUC and AUPRC of 50 experiments for each method and perform the method comparison using two-sided paired Wilcoxon rank-sum test. Since standardizing input data is a common practice for deep learning approach, we further compare the performance using unscaled beta values and scaled beta values with mean 0 and variance 1.

When the beta values is unscaled (Figure 2A), for predicting CN-to-MCI progression, MT-CAE and MT-LSTMAE are top-ranked methods in terms of median AUC (MT-CAE: 0.996, MT-LSTMAE: 0.991) followed by LSTM1, CNN1, CNNLSTM1 and LSTM2 (LSTM1: 0.900; CNN1: 0.893; CNNLSTM1: 0.893; LSTM2: 0.891). We also find that the the improvement of MT-CAE and MT-LSTMAE over third-ranked LSTM1 is significant (MT-CAE vs LSTM1: pvalue=1.923×10^−17^; MT-LSTMAE vs LSTM1: pvalue=3.085×10^−15^) based on the two-sided Wilcoxon rank sum test on 50 AUC values. The improvement of multi-task autoencoders may be attributed to the advantage of multi-task learning, which can learn the compressed feature representations of high-dimensional methylation values and utilize the compressed feature representations for the prediction simultaneously. In other words, the learnt feature representations not only capture as much as information as possible from the methylation data in the reconstruction but are also tailored to improve the prediction performance. Interestingly, other deep learning models hold a comparable performance (AUC is between 0.8 to 0.9) except for CNNLSTM2 (AUC=0.704). Moreover, the prediction performance of random forest is close to random guess (AUC=0.483).

**Figure 2:**
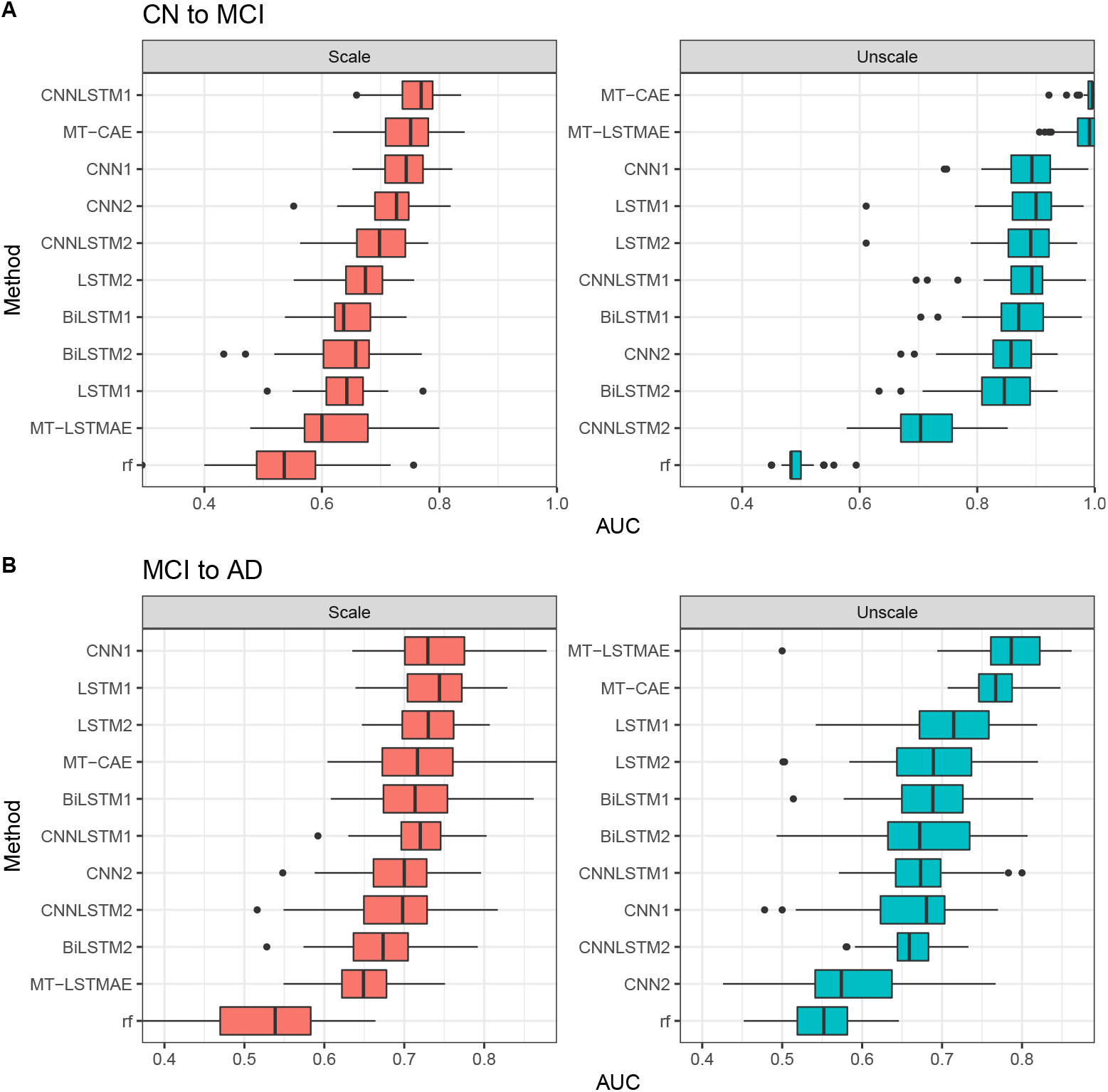
Compare proposed multi-task deep autoencoders (MT-CAE and MT-LSTMAE) and competing machine learning methods in terms of AUC for (A) CN-to-MCI (B) MCI-to-AD

Moreover, by evaluating compared models in different categories, we have additional observations on the impact of network architecture on the model performance. First, considering moderate size of training samples, increasing number of layer, either LSTM layer or convolutional layer, does not necessarily improve the performance, which is evident by comparing CNN1 and CNN2 (0.893 vs 0.858), LSTM1 and LSTM2 (0.900 vs 0.891), BiLSTM1 and BiLSTM2 (0.871 vs 0.846), and CNNLSTM1 and CNNLSTM2 (0.893 vs 0.704). More layers may even result in overfitting such as CNNLSTM2. Second, adding backwards direction of information flow, does not lead to the improvement of performance by comparing LSTM1 and BiLSTM1 (0.900 vs 0.871), and LSTM2 and BiLSTM2 (0.891 vs 0.846). Third, deep learning approaches significantly outperforms random forest.

When using scaled beta values as model input, we find that MT-CAE together with single-task deep learning models such as CNNLSTM1, CNN1 and CNN2 have the best performance. However, the performance of most approaches decrease dramatically. Notably, the median AUC of MT-LSTMAE drops mostly from 0.991 to 0.600 (39.1%), and the median AUC of MT-CAE drops from 0.996 to 0.751 (24.5%). Comparing LSTM/BiLSTM and CNN, the median AUC of LSTM drops more than that of CNN (LSTM1 from 0.900 to 0.643 (25.7%) and LSTM2 from 0.891 to 0.674 (21.7%); BiLSTM1 from 0.871 to 0.637 (23.4%) and BiLSTM2 from 0.846 to 0.658 (18.8%); CNN1 from 0.893 to 0.744 (14.9%) and CNN2 from 0.858 to 0.727 (13.1%)). The declined performance of a hybrid of CNN and LSTM such as CNNLSTM1 (from 0.893 to 0.769 (12.4%)) is also mild compared to LSTM1. However, we find the performance of CNNLSTM2 remains stable (from 0.704 to 0.698) and the median AUC of random forest slightly increases from 0.483 to 0.536.

For predicting MCI-to-AD progression, we have the similar findings as CN-to-MCI progression. When the beta values are unscaled, MT-LSTMAE and MT-CAE achieve the top performance (MT-LSTMAE: 0.787, MT-CAE: 0.767), followed by LSTM1 (0.715). The improvement is also significant (MT-LSTMAE vs LSTM1: pvalue=1.882×10^−8^; MT-CAE vs LSTM1: pvalue=2.707×10^−6^) based on the two-sided Wilcoxon rank sum test on 50 AUC values. Other deep learning models hold a comparable performance with the AUC between 0.6 to 0.7 except for CNN2 (AUC=0.574). Moreover, the prediction performance of random forest is slightly better than random guess (AUC=0.552). When the beta values are scaled, the AUC of MT-LSTMAE has the most significant drop from 0.787 to 0.649 (17.48%). In contrast, MT-CAE has a slight decline from 0.767 to 0.717 (6.584%). Other deep learning approaches remain stable except CNN2 with a significant increased AUC from 0.574 to 0.700 (21.95%).

Besides AUC, we also examine the performance in terms of AUPRC (Figure S1). The overall trend is similar to AUC. Overall, these findings indicate that using longitudinal methylation data in peripheral blood can accurately predict AD progression for both CN-to-MCI conversion and MCI-to-AD conversion. Moreover, the proposed multi-task autoencoders MT-CAE and MT-LSTMAE significantly improve the prediction performance especially when using unscaled beta values. However, the performance of MT-LSTMAE declines significantly when using scaled beta values. Compared to MT-LSTMAE, MT-CAE is less sensitive to the data transformation of beta values. For most approaches, unscaled beta values is more favorable than scaled beta values as the model input especially for predicting CN-to-MCI conversion. Moreover, random forest has the poorest performance in both conversions.

### 3.2 Multi-task deep autoencoders improve the prediction performance than single-task deep autoencoders

In the previous section, we’ve demonstrated that proposed two multi-task deep autoencoders, MT-CAE and MT-LSTMAE, improve the prediction for AD progression using temporal DNA methylation data compared to classic deep learning approaches. In this section, we will evaluate the advantage of multi-task autoencoder compared to single-task autoencoder. Different from multi-task autoencoder, single-task autoencoder performs the reconstruction and prediction in two sequential steps. Specifically, we investigate three single-task autoencoders: (i) convolutional autoencoder (CAE), which has the same network architecture of autoencoder as that of MT-CAE but with one task for minimizing the reconstruction error to learn the compressed feature representations. Then, the compressed feature representations will be sent to the same network architecture of feedforward neural network as that of MT-CAE for predicting AD progression; (ii) LSTM autoencoder (LSTMAE), which has the same network architecture of autoencoder as that of MT-LSTMAE with one task for minimizing the reconstruction error to learn the compressed feature representations. Then, the compressed feature representations will be sent to the same network architecture of feedforward neural network as that of MT-LSTMAE for predicting AD progression; (iii) a standard autoencoder (AE) using three dense layers for both encoder and decoder. Since dense layers is not feasible to take temporal data, we train the standard autoencoder for each time step to learn the compressed feature representation for each time step. The compressed feature representations for all time steps are further merged in a temporal format, which will be sent to a LSTM network for predicting AD progression. We denote the two-step approach as AE-LSTM.

We use the same strategy to create training, testing and validation datasets and repeat the experiment 50 times for predicting CN-to-MCI and MCI-to-AD progression respectively. We further evaluate the performance of all models using both unscaled or scaled beta values. We report the AUC and AUPRC from 50 experiments for each method and perform the method comparison using two-sided paired Wilcoxon rank-sum test (Figure 3). Consequently, when using unscaled beta values to predict CN-to-MCI progression, we find that MT-LSTMAE significantly outperforms its single-task counterpart LSTMAE in terms of median AUC (0.991 vs 0.580, pvalue=6.229×10^−18^). Similarly, MT-CAE also holds a clear advantage to its single-task counterpart CAE in terms of median AUC (0.996 vs 0.680, pvalue=5.173×10^−18^). Moreover, both multi-task deep autoencoders and their single-task counterparts perform much better than AE-LSTM (AUC=0.659). The worst performance of AE-LSTM may be explained by the temporal information is disrupted when an independent AE is developed to learn compressed feature representations for each time step. When using scaled beta values, the performance of both MT-CAE and MT-LSTMAE decline. However, the above trends still hold. MT-CAE has the best performance, followed by MT-LSTMAE and their single-task counterparts. Again, AE-LSTM has the worst performance. Notably, the AUC of AE-LSTM drops dramatically from 0.648 to 0.555 (9.3%). In contrast, CAE and LSTMAE remains very stable (CAE: 0.680 vs 0.620; LSTMAE: 0.580 vs 0.563).

**Figure 3:**
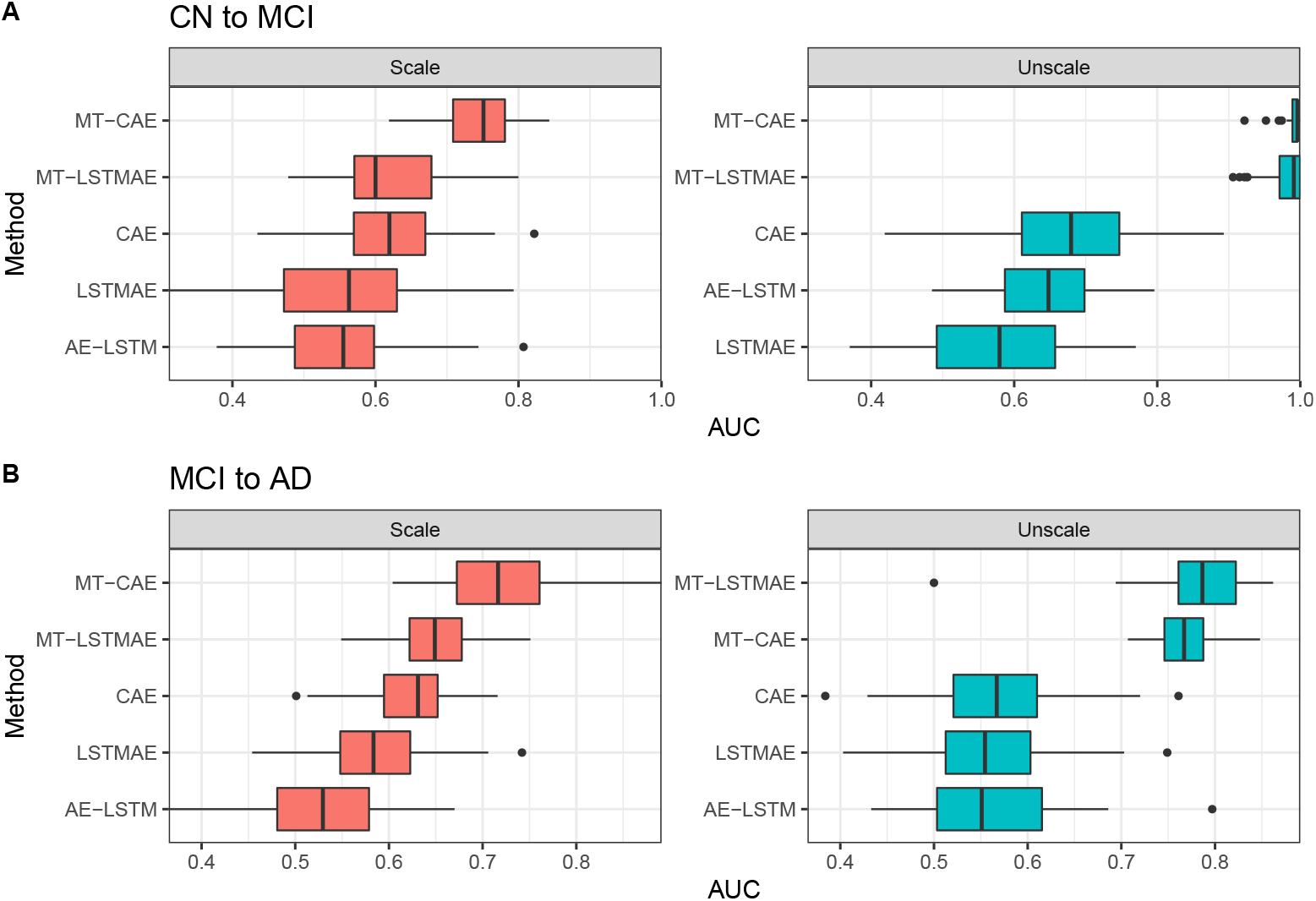
Compare multi-task deep autoencoders and single-task counterparts in terms of AUC. Specifically, we investigate three single-task autoencoders: (i) convolutional autoencoder (CAE), which has the same network architecture of autoencoder as that of MT-CAE but with one task for minimizing the reconstruction error to learn the compressed feature representations. Then, the compressed feature representations will be sent to the same network architecture of feedforward neural network as that of MT-CAE for predicting AD progression; (ii) LSTM autoencoder (LSTMAE), which has the same network architecture of autoencoder as that of MT-LSTMAE with one task for minimizing the reconstruction error to learn the compressed feature representations. Then, the compressed feature representations will be sent to the same network architecture of feedforward neural network as that of MT-LSTMAE for predicting AD progression; (iii) a standard autoencoder (AE) using three dense layers for both encoder and decoder. Since dense layers is not feasible to take temporal data, we train the standard autoencoder for each time step to learn the compressed feature representation for each time step. The compressed feature representations for all time steps are further merged in a temporal format, which will be sent to a LSTM network for predicting AD progression. We denote the two-step approach as AE-LSTM.

For predicting MCI-to-AD progression, we find the overall trend is similar to that of predicting CN-to-MCI progression. MT-LSTMAE and MT-CAE top the performance either using the scaled or unscaled beta values. When using scaled beta values, the performance of both multi-task models decline, while MT-CAE is more robust than MT-LSTMAE. Still, AE-LSTM has the poorest performance. Moreover, the performance of all single task autoencoders remain stable despite the data transformation of beta values.

Additionally, we report the performance in terms of AUPRC and find a similar trend (Figure S2). Overall, the above findings indicate that multi-task deep autoencoders hold a clear advantage than its single-task counterparts despite the input data transformation. Moreover, a common practice of two-step approach, which using autoencoder for feature extraction and an independent classifier using extracted features for prediction, has unfavorable performance.

### 3.3 Temporal information improves the prediction for AD progression

In the previous sections, all temporal methylation data from time 1 to *T* has been used to predict AD progression at time *T*. Here, we are interested in exploring (i) whether using historical data (e.g. 1 to *T* − 1) only can accurately predict AD progression at time *T* ; (ii) whether using all temporal data improves the prediction for AD progression than using historical data only. Accordingly, we use both temporal and historical data as model input for two proposed multi-task deep autoencoders. As a comparison, we also include aforementioned deep learning models, which are single-task models and do not utilize autoencoder for feature extraction. Since increasing number of layers is not helpful in improving the performance, we only include the shallow counterpart for each compared deep learning model (Table 1).

When using unscaled beta values, we find that using historical methylation data can accurately predict CN-to-MCI progression (Figure 4A). Two proposed multi-task models (MT-CAE 1t: 0.954, MT-LSTM 1t: 0.956) rank top among all models using historical methylation data only. Moreover, using historical data only decreases the performance in terms of AUC for both MT-CAE (MT-CAE: 0.996 vs MT-CAE 1t: 0.954; pvalue= 9.486×10^−15^) and MT-LSTMAE (MT-LSTMAE: 0.991 vs MT-LSTM 1t: 0.956; pvalue=6.055×10^−8^). The same trend holds for other compared deep learning approaches, which do not involve feature extraction using autoencoder. For example, the median AUC decreases from 0.893 to 0.811 (pvalue=2.34×10^−11^) for CNN1, from 0.900 to 0.826 (pvalue=1.929×10^−9^) for LSTM1, from 0.871 to 0.819 (pvalue=2.752×10^−5^) for BiLSTM1, and from 0.893 to 0.815 (pvalue=4.298×10^−11^) for CNNLSTM1. Notably, we find the performance of compared deep learning models decline more compared to multi-task autoencoders (MT-CAE: 4.2%; MT-LSTMAE: 3.5% compared to CNN1: 8.2%; LSTM1: 7.4%; BiLSTM1: 5.2%; CNNLSTM1: 7.8%). A similar trend is found when using scaled beta values, that is, using historical methylation data can achieve an accurate prediction but will deteriorate the performance compared to using all temporal methylation data, and the performance of other deep learning models decline more compared to multi-task autoencoders. For MCI-to-AD progression, we find the global trend is similar to that of CN-to-MCI progression (Figure 4B).

**Figure 4:**
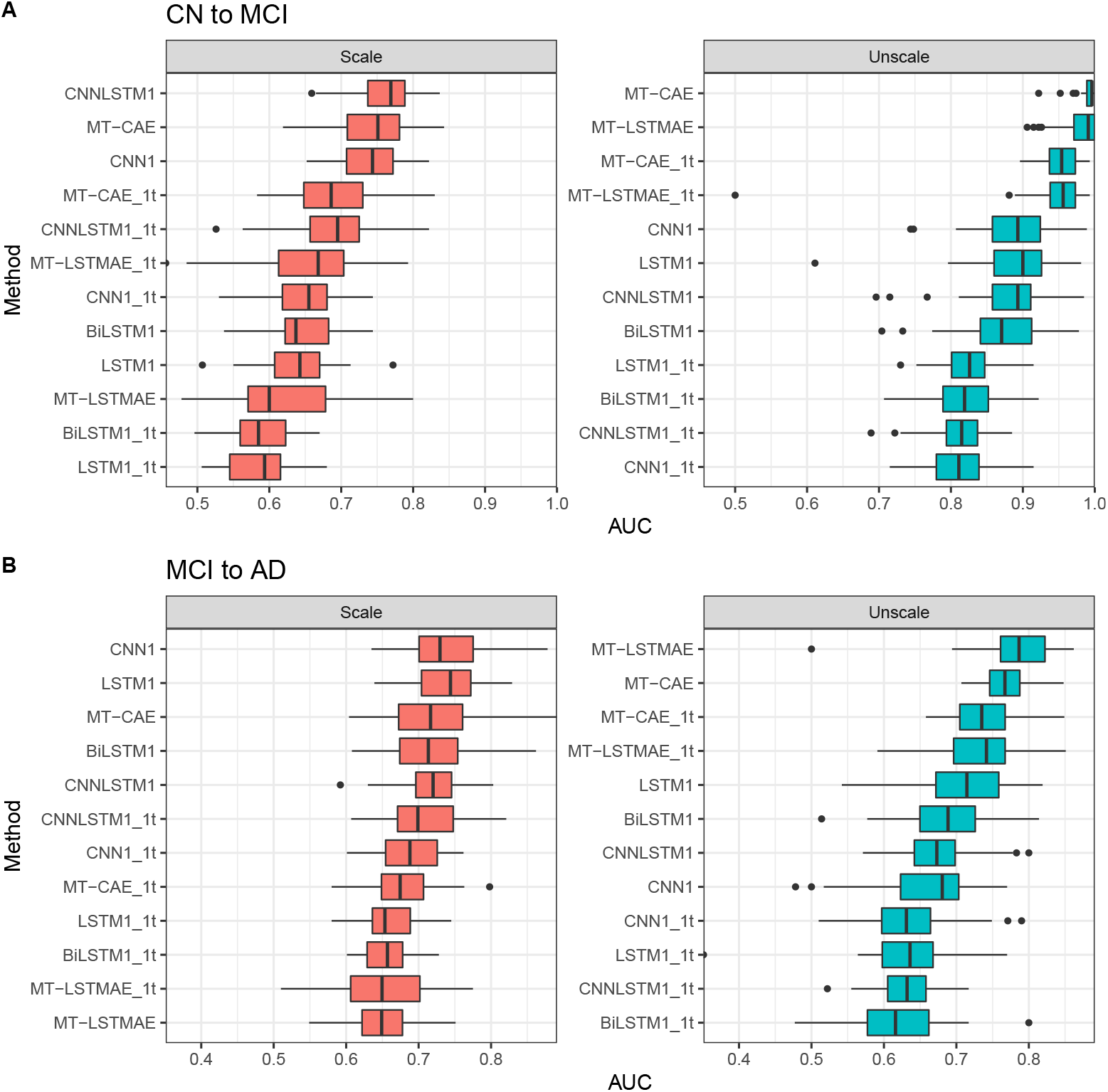
Compare using historical methylation data and all temporal methylation data for predicting AD progression in terms of AUC. MT-CAE 1t: MT-CAE uses only methylation data at first time step; MT-LSTMAE 1t: MT-LSTMAE uses only methylation data at first time step; CNN1 1t: CNN1 uses only methylation data at first time step; LSTM1 1t: LSTM1 uses only methylation data at first time step; BiL-STM1 1t: BiLSTM1 uses only methylation data at first time step; CNNLSTM1 1t: CNNLSTM1 uses only methylation data at first time step.

In addition, we report the performance in terms of AUPRC for all methods and find similar observations (Figure S3). Overall, the above observations indicate that despite input data transformation, using historical methylation data alone can predict the AD progression accurately, and including all temporal methylation data can further improve the prediction for both proposed multi-task deep autoencoders and compared deep learning approaches, which do not adopt autoencoder for feature extraction.

### 3.4 Multi-task deep autoencoder improves the reconstruction for temporal methylation data

Both MT-CAE and MT-LSTMAE are multi-task deep autoencoders, which have with the primary task for predicting AD progression and auxiliary task for reconstructing temporal methylation data. Here, we will evaluate the performance for the auxiliary task and compare multi-task models with a standard autoencoder (AE). For AE, we train an independent three-dense-layer autoencoder to reconstruct the methylation data for each time step.

Same as the prediction task, in the reconstruction task, 20% of the total samples is used as independent testing, 80% is used as training set among which 20% is used as validation set. To control the bias from random sampling, the aforementioned procedure is repeated 50 times. We use sample-wise Pearson correlation (R) and mean square error (MSE) two metrics to measure the reconstruction performance. Specifically, R is calculated between observed and reconstructed beta values for each sample in each time step. MSE is calculated between observed and reconstructed beta values for all samples in all time steps. We report the R and MSE of 50 experiments for each method and perform the method comparison using two-sided paired Wilcoxon rank-sum test.

For CN-to-MCI progression (Figure 5A), for scaled beta values, we find that multi-task deep autoen-coders obtain lower MSE (MT-CAE: 0.908; MT-LSTMAE: 0.838) than AE (0.914) and higher R (MT-CAE: 0.254; MT-LSTMAE: 0.336) than AE (0.246). Moreover, the improvement of MT-LSTMAE over AE is significant by reducing MSE 7.6% (pvalue*<*2.2×10^−16^) and increasing R by 9% (pvalue*<*2.2×10^−16^). For unscaled beta values, the performance of all methods improve significantly. MSE decreases from 0.908 to 0.026 by 88.2% for MT-CAE, from 0.838 to 0.003 by 83.5% for MT-LSTMAE and from 0.914 to 0.143 by 77.1% for AE. Accordingly, R increases from 0.254 to 0.938 by 68.4% for MT-CAE, 0.336 to 0.992 by 65.6% for MT-LSTMAE and 0.246 to 0.689 by 44.3% for AE. Still, MT-CAE and MT-LSTMAE have a clear advantage over AE in terms of both lower MSE (MT-LSTMAE: 0.003; MT-CAE: 0.026; AE: 0.143) and higher R (MT-LSTMAE 0.992; MT-CAE: 0.938; AE: 0.689). MT-LSTMAE has the best performance by obtaining the highest R and lowest MSE. Notably, the performance of MT-CAE and MT-LSTMAE is more significantly improved than AE.

**Figure 5:**
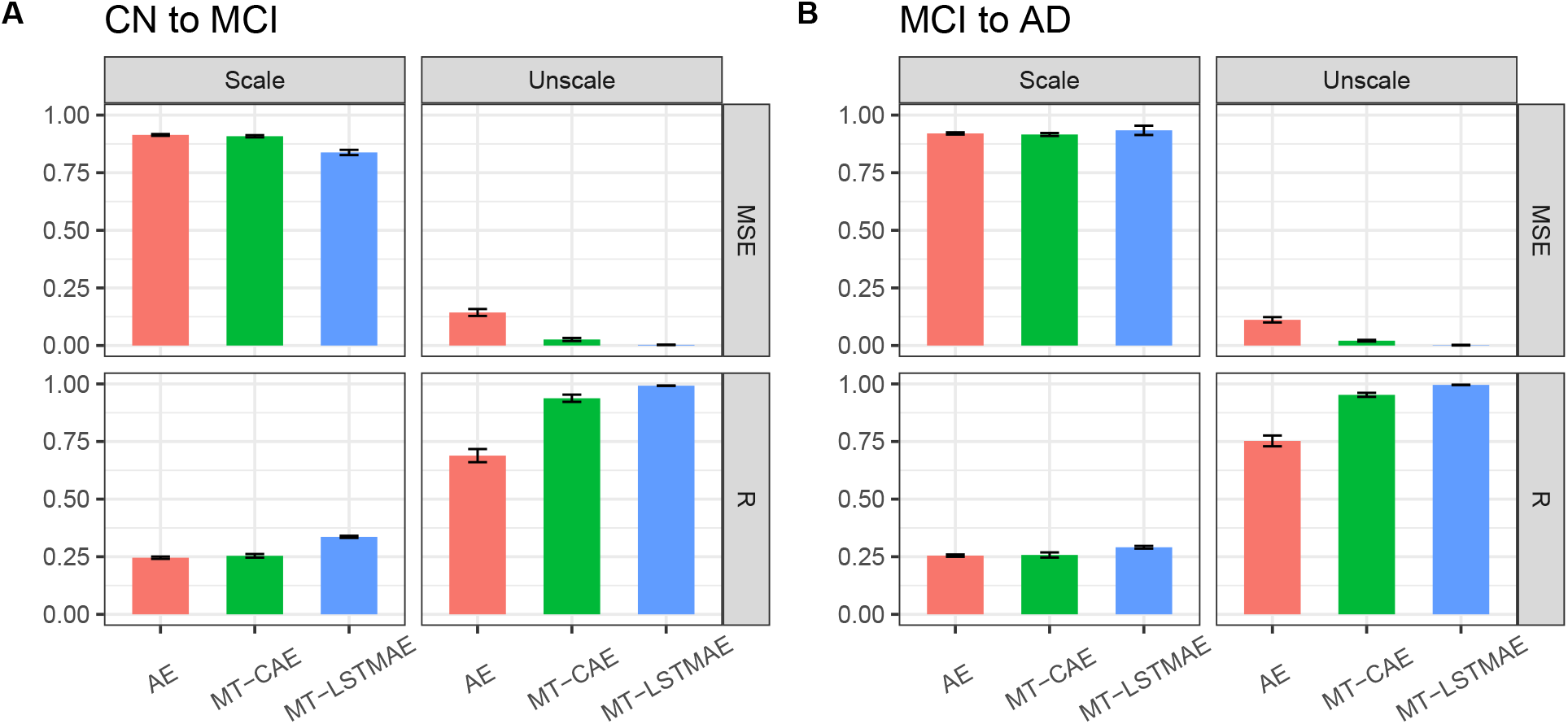
Compare MT-LSTMAE, MT-CAE and AE for reconstructing both scaled and unscaled beta values in CN-to-MCI (A) and MCI-to-AD (B).

For MCI-to-AD progression (Figure 5B), for scaled beta values, the three approaches have comparable performance in terms of (R MT-LSTMAE: 0.291; MT-CAE: 0.258; AE: 0.255) and MSE (MT-LSTMAE: 0.934; MT-CAE: 0.916; AE: 0.920). Similar to CN-to-MCI progression, the performance of all methods improve significantly for unscaled beta values. Both MT-LSTMAE and MT-CAE outperform AE by achieving a higher R (MT-LSTMAE: 0.996; MT-CAE: 0.953; AE: 0.753) and lower MSE (MT-LSTMAE: 0.002; MT-CAE: 0.021; AE: 0.112). Notably, MT-LSTMAE has the best performance by achieving the highest R and lowest MSE.

Overall, we find that proposed multi-task deep autoencoders MT-LSTMAE and MT-CAE have an advantage over the standard autoencoder AE for reconstructing methylation data, which are either represented by scaled or unscaled beta values. The advantage is more evident when the beta values are unscaled. Particularly, MT-LSTMAE has the best performance. In addition, using unscale beta values achieves much better reconstruction results in terms of both MSE and R for all approaches, which indicates that unscaled beta values is a better choice for performing DNA methylation imputation.

### 3.5 Sensitivity analysis of feature size on the performance of multi-task deep autoencoder

**Figure 6:**
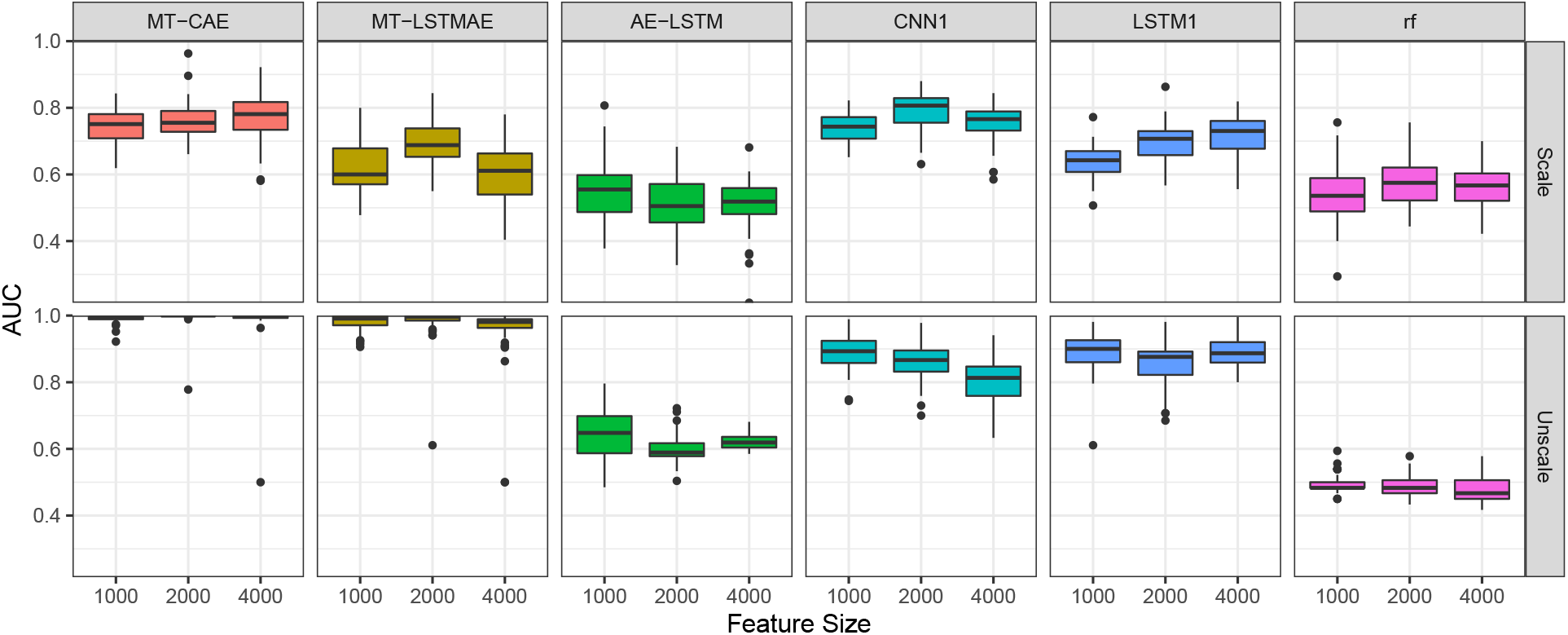
Comparisons of prediction performance for multi-task deep autoencoders (MT-CAE and MT-LSTMAE), single-task autoencoder with LSTM as the classifier (AE-LSTM), deep learning models not based on autoencoder (CNN1 and LSTM1) and non-deep learning approach (random forest). 1000 to 2000 and 4000 most informative CpGs are selected as input methylation features.

In the above sections, we use proposed feature selection approach to choose 1000 most informative CpGs as input methylation features for developing and evaluating all models. Here, we will perform a sensitivity analysis to evaluate whether 1000 most informative CpGs are sufficient for proposed multi-task deep autoencoders and competing approaches. To do this, we use the same feature selection approach to choose 2000 and 4000 most informative CpGs as input methylation features and perform the same experiments for all methods. Without loss of generality, we evaluate two multi-task deep autoencoders (MT-CAE and MT-LSTMAE), single-task autoencoder with LSTM as the classifier (AE-LSTM), deep learning models not based on autoencoder (CNN1 and LSTM1) and non-deep learning approach (random forest) on the CN-to-MCI progression. As aforementioned, the experiments are repeated 50 times and the 50 AUCs are reported.

Consequently (Figure6), for scaled beta values, we find that the median AUC of MT-CAE remain stable despite the change of feature size (1000: 0.751; 2000: 0.755; 4000: 0.781). The stable AUC can also be found for AE-LSTM (1000: 0.555; 2000: 0.505; 4000: 0.519) and random forest (1000: 0.536; 2000: 0.575; 4000: 0.567). The AUC of LSTM1 has a steady increase from 0.643 to 0.707 to 0.730. For MT-LSTMAE and CNN1, the AUC increases at 2000 CpGs but decline at 4000 CpGs. For unscaled beta values, the median AUC of MT-CAE and MT-LSTMAE remain stable across different feature size (MT-CAE 1000: 0.996; 2000: 1.000; 4000: 1.000; MT-LSTMAE 1000: 0.991; 2000: 0.996; 4000: 0.981). Again, the stable AUC can also be found for random forest (1000: 0.483; 2000: 0.483; 4000: 0.467). However, the median AUC of AE-LSTM is maximum when the feature size is 1000 (1000: 0.648; 2000: 0.589; 4000: 0.619). Similar to AE-LSTM, the median AUC of CNN1 declines steadily with the increase of feature size (1000: 0.893; 2000: 0.867; 4000: 0.813). Interestingly, LSTM1 has a declined AUC at 2000 CpGs but increased AUC at 4000 CpGs with a comparable AUC between 1000 and 4000 CpGs (1000: 0.900; 2000: 0.876; 4000: 0.887).

Overall, we find that increasing the feature size has a limited impact on the prediction performance for all approaches especially when using unscaled beta values. Since using unscaled beta values improves prediction performance for all methods, and expanding feature size will increase the model training time and consume more computer memory, 1000 most informative CpGs are sufficient for developing prediction models in this study.

## Discussion

Traditional approaches such as imaging and cerebrospinal fluid for AD diagnosis are the invasive or expensive. These limitations make the biomarker obtained from peripheral tissues, e.g. blood, a potentially useful diagnostic tool due to its favorable noninvasive and easily accessible characteristics. However, whether DNA methylation data in peripheral blood can be utilized for AD diagnosis especially in a longitudinal study is seldom exploited.

In this work, we develop two multi-task deep autoencoders, which has the primary task for predicting AD progression in a longitudinal study and an auxiliary task for reconstructing temporal DNA methylation data simultaneously. The first multi-task deep autoencoder named MT-CAE is based on convolutional autoencoder. The encoder consists of convolutional layers and the decoder consists of transposed convolutional layers. By taking the advantage of convolution operation, MT-CAE can capturing both the spatial and temporal information of longitudinal methylation data. Besides being sent to the decoder for the reconstruction task, the output of the encoder, which is known as compressed feature representations, connects to a feed-forward feedforward network for the prediction task. It should be noted that the dimension of feature representations is largely reduced compared to input methylation features. The second multi-task autoencoder named MT-LSTME is based on LSTM autoencoder. Specifically, the encoder is composed of two-level stacked LSTM layers to learn the high-order nonlinear temporal dependency of longitudinal methylation data. Different from the encoder, the decoder consists of one LSTM layer with *T* LSTM units, and each LSTM unit is followed by a dense layer with shared weights across *T* time steps. Similar to MT-CAE, the output of the encoder, which is the compressed feature representations, is treated as model input to the feed-forward feedforward network classifier.

Different from a typical way for high-dimensional data prediction, which utilize a single-task autoencoder for compressed feature extraction followed by a classifier (e.g. CNN, LSTM) with the extracted features as model input in two separate steps, both feature extraction and classification are performed simultaneously in multi-task deep autoencoders, which offers the advantage to reduce overfitting through shared representations. Specifically, the prediction task and reconstruction task share all parameters in the encoder for multi-task deep autoencoders. We further develop a hybrid loss function, which is a weighted average of prediction loss and reconstruction loss. By minimizing the hybrid loss function, the multi-task deep autoencoders aim to improve the learning for the compressed feature representations by considering the accuracy of both prediction and reconstruction.

To demonstrate the advantage of proposed multi-task deep autoencoders, we include classic deep learning approaches for time-series classification such as convolutional neural networks, LSTM network and a hybrid of CNN and LSTM network as well as random forest, which has been frequently used for high-dimensional data prediction. We have two important findings. First, MT-CAE and MT-LSTMAE significantly outperform other deep learning methods in predicting CN-to-MCI and MCI-to-AD conversions when the beta values are unscaled. Notably, compared to MT-LSTMAE, MT-CAE is less sensitive to the data transformation of beta values. Instead, random forest, which is a non-deep learning method, has the poorest performance. Second, unscaled beta values are more favorable compared to scaled beta values as the model input for the prediction task, as it improves the overall performance for most models especially for multi-task deep autoencoders. These results indicate that proposed multi-task deep autoencoders hold a clear advantage to classic deep learning approaches to predict AD progression by using longitudinal DNA methylation data in peripheral blood.

To demonstrate the superiority of multi-task learning to single-task learning, we also include the two-step approach as the comparison. We evaluate three autoencoders for feature extraction, which include (i) a single-task convolutional autoencoder (CAE) with the same network architecture of autoencoder as that of MT-CAE to learn the compressed feature representations, which is sent to the same classifier of MT-CAE; a single-task LSTM autoencoder (LSTMAE) with the same network architecture of autoencoder as that of MT-LSTMAE to learn the compressed feature representations, which is sent to the same classifier of MT-LSTMAE; (iii) a standard autoencoder (AE) to learn the compressed feature representation for each time step, which are further merged to a LSTM classifier (AE-LSTM). Consequently, MT-CAE and MT-LSTMAE significantly outperform their single-task counterparts and AE-LSTM despite the transformation of beta values. AE-LSTM has the worst performance. These observations validate the advantage of multi-task learning by jointly modeling the prediction and reconstruction. Furthermore, we demonstrate that using historical methylation data can achieve an accurate prediction for AD progression but will have a decline of the performance compared to using all temporal methylation data. Notably, the performance of classic deep learning models decline more compared to proposed multi-task deep autoencoders.

Lastly, we evaluate the performance of reconstructing temporal methylation data and compare multi-task deep autoencoders to standard autoencoder. Consequently, multi-task deep autoencoders improve the reconstruction for temporal methylation data, which is measured by the higher correlation between observed and reconstructed methylation profile on the individual level, as well as the smaller difference between observed and reconstructed methylation profile. MT-LSTMAE has the overall best performance. In addition, though the performance of all models improves when using unscaled beta values, the improvement of multi-task models is more evident. Overall, joint modeling both feature extraction and prediction improve the performance for both prediction and reconstruction due to the benefit of multi-task learning. Moreover, using unscaled beta values not only improves the prediction performance but also improves the reconstruction performance.

To create the positive set in the classifier, we consider two types of AD conversion: CN-to-MCI and MCI-to-AD because they are the majority in the data collection in ADNI. Accordingly, non-conversion groups such as CN-to-CN and MCI-to-MCI are treated as the negative sets. The number of visits for each individual with DNA methylation profiled are uneven in the ADNI, which ranges from minimum one and maximum five. However, we only include methylation profiles from two time steps for each individual, which are the first visit and last visit as the training features for both positive and negative sets. The rationale lies on three aspects. First, in most cases, the diagnosis for the first visit is the baseline status (e.g. CN) and diagnosis for the last visit is the converted status (e.g. MCI). Therefore, the profiled methylation data in the first visit and last visit reflect the AD progression. Second, including first and last visits maximizes the inclusion of samples. Third, though some methods for missing data imputation can impute the methylation profiles for missed visits, they may suffer from the infeasibility to accurately impute high-dimensional methylation data. In addition, imputed methylation data may be deviated from the truth and will bias the results. Indeed, using historical data only, i.e. methylation data in the first visit, still achieves desirable prediction performance. This empirical evidence further validates that it is appropriate to use two time points in the longitudinal study for predicting AD progression.

There are limitation for this work. First, for longitudinal methylation data available in ADNI, the sample size and number of time steps are still small. With the further reduction in sequencing costs, we anticipate a large-scale longitudinal DNA methylation data in both sample size and number of visits will be available in the near future to further improve the model performance. Second, we are only interested in evaluating whether DNA methylation data in peripheral blood is able to predict AD progression. To further improve the prediction performance, the proposed deep learning models can be extended to integrate other types of omics data in peripheral blood, which include but not limited to transcriptome, proteome and metabolome data. We will extend the multi-task deep autoencoders into a multi-modal multi-task deep learning framework to handle heterogeneous multi-omics input when they are available.

## Data Availability

All data produced are available online at https://adni.loni.usc.edu/

## Acknowledgement

This work was supported by Indiana University Precision Health Initiative, Showalter Research Trust and National Institute of General Medical Sciences of the National Institutes of Health under Award Number R35GM142701 to LC.

